# A quantitative evaluation of the impact of vaccine roll-out rate and coverage on reducing deaths from COVID-19: a counterfactual study on the impact of the delayed vaccination programme in Iran

**DOI:** 10.1101/2023.05.31.23290799

**Authors:** Mahan Ghafari, Sepanta Hosseinpour, Mohammad Saeid Rezaee-Zavareh, Stefan Dascalu, Somayeh Rostamian, Kiarash Aramesh, Kaveh Madani, Shahram Kordasti

## Abstract

Vaccination has been a crucial factor in the fight against COVID-19 because of its effectiveness in suppressing virus circulation, lowering the risk of severe disease, and ultimately saving lives. Many countries with an early and rapid distribution of COVID-19 vaccines performed much better in reducing their total number of deaths than those with lower coverage and slower roll-out pace. However, we still do not know how many more deaths could have been averted if countries with slower vaccine roll-outs followed the same rate as countries with earlier and faster distribution of vaccines. Here, we investigated counterfactual scenarios for the number of avertable COVID-19 deaths in a given country based on other countries’ vaccine roll-out rates. As a case study, we compared Iran to eight model countries with similar income brackets and dominant COVID-19 vaccine types. Our analysis revealed that faster roll-outs were associated with higher numbers of averted deaths. While Iran’s percentage of fully vaccinated individuals would have been similar to Bangladesh, Nepal, Sri Lanka, and Turkey under counterfactual roll-out rates, adopting Turkey’s rates could have averted up to 50,000 more deaths, whereas following Bangladesh’s rates could have led to up to 52,800 additional losses of lives in Iran. Notably, a counterfactual scenario based on Argentina’s early but slow roll-out rate resulted in a smaller number of averted deaths in Iran, up to 12,600 more individuals. Following Montenegro’s or Bolivia’s model of faster per capita roll-out rates for Iran could have resulted in more averted deaths in older age groups, particularly during the Alpha and Delta waves, despite their lower overall coverage. Also, following Bahrain’s model as an upper bound benchmark, Iran could have averted 75,300 deaths throughout the pandemic, primarily in the >50 age groups. This study provides insights into future decisions on the management of infectious disease epidemics through vaccination strategies by comparing the relative performance of different countries in terms of their timing, pace, and coverage of vaccination in preventing COVID-19 deaths.

## Introduction

On 4 May, 2023, the World Health Organization (WHO) determined that the public health emergency phase of the COVID-19 pandemic was over and laid out recommendations for countries on their transition into the long-term management of COVID-19, among other infectious diseases [1]. While the announcement that the healthcare emergency was declared over may have brought some relief to the public, it did not signify an end to the pandemic. As such, governments still need to stay vigilant and support the global effort to take necessary actions for suppressing COVID-19 transmission and reducing the burden of the disease. Among the various public health measures that were implemented throughout the pandemic, maximising the impact of vaccination worldwide has been a key factor in bringing the public health emergency to an end [2]. In this respect, lessons derived from national COVID-19 vaccination campaigns worldwide are crucial as they demonstrate the successes and shortcomings of different countries in reducing the burden of disease during a public health emergency.

Since early 2021, vaccination has been a major contributor to reducing the burden of COVID-19 globally [3, 4]. However, its benefits were not equitably distributed to every country partly due to challenges with the production, distribution, and affordability of vaccines [5, 6] despite efforts to fairly allocate them globally [7] and partly due to vaccine hesitancy [8], which remains a threat to global health. While many studies have focused on quantifying the effectiveness of vaccination campaigns through estimating the number of prevented deaths as a result of vaccination [3, 9–11], fewer studies focused on quantifying the impact of national vaccination programmes’ speed and timings on reducing the burden of COVID-19 [12–15]. More specifically, very limited attention has been given to quantitatively comparing countries’ relative performance based on their vaccine roll-out rates on reducing the burden of COVID-19.

While several countries initiated the process of purchasing vaccines towards the end of 2020 and early 2021 through local production, bilateral advance purchase agreements, and the COVID-19 Vaccine Global Access Facility (COVAX) [16, 17], Iran faced several challenges with securing an adequate and timely supply of vaccines. This was partly due to limited global vaccine production capacity and high demand, and in part due to geopolitical factors that were unique to Iran [18]. A prime example of geopolitical tensions was the decision to ban the importation of vaccines from the US and UK to Iran [19]. The country also was not on the first interim distribution forecast list from COVAX and received its first batch on 5 April 2021 [20] while several other countries received them nearly a month earlier [21, 22].

These factors, along with several others such as delayed home-grown production of vaccines and importations from other countries contributed to a delayed start of the vaccination campaign in Iran. As a result, six months after the start of their vaccination, only less than 5% of the Iranian population were fully vaccinated [18]. Evaluating Iran’s vaccination campaign provides an opportunity to understand the impact of delayed vaccination on reducing the number of vaccine-preventable deaths. It can also shed light on the factors that contributed to the increased burden of COVID-19 relative to countries with faster vaccination roll-out and identify potential problems and best practices in vaccination strategies that can be applied more broadly to other countries in the future.

In this work, we developed a framework to retroactively calculate the age-stratified number of avertable deaths in Iran had the country followed the same per capita roll-out rates as other countries. By selecting eight model countries that predominantly vaccinated their populations with inactivated virus vaccines, we examined the impact of vaccination programme start dates, roll-out rates, and overall coverage on avertable COVID-19 deaths (ACDs) in Iran.

## Results

With nearly three-quarters of its eligible population fully vaccinated against COVID-19 by late April 2022, Iran had a relatively high vaccine coverage compared to many other upper- and lower-middle-income countries [23]. However, despite its high coverage, it under-performed compared to almost all other LMICs and UMICs with similar coverages, ranking 29th out of all 44 UMICs, as determined by the number of deaths averted per person by December 2021 (**Supplementary Figure 1**) [3].

To investigate the extent to which Iran’s performance could have been enhanced or diminished if alternative vaccination strategies were employed, we selected eight model countries from other upper- and lower-middle-income countries, with the exception of Bahrain (see **Methods**), a high-income country, that similar to Iran predominantly used inactivated virus vaccines particularly the BBIBP-CorV vaccine (see **Supplementary Table 1** and **Supplementary Table 2**). We then examined counterfactual scenarios whereby Iran’s vaccine roll-out rate is replaced with that of the model countries to see if Iran would have had more (or fewer) ACDs (see **Methods**).

Our findings showed that for a fixed overall percentage of fully vaccinated individuals, faster roll-out rates are associated with higher ACDs. While the percentage of fully vaccinated Iranians based on a counterfactual vaccine roll-out rate from Bangladesh, Nepal, Sri Lanka, and Turkey would have roughly been the same (∼65-70%), Iran could have averted as many as 50,000 (95% confidence interval: 38,100 - 53,500) deaths if it had followed Turkey’s per capita roll-out rates and would have had as many as 52,800 (17,400-189,500) more deaths if it had followed Bangladesh (**Table 1**). This is because Turkey started its vaccination programme two months earlier than Bangladesh (and Iran) and also had a much faster per capita roll-out rate. On the other hand, following Argentina’s per capita roll-out rates, Iran would have only averted as many as 12,600 (95% CI: 10,400 - 13,300) deaths despite the fact that Argentina, similar to Turkey, had an early vaccination start date. This is because Argentina had a much slower roll-out rate and, as a result, fewer deaths would have been averted during the Alpha and Delta waves in Iran, particularly in the older age groups (**Table 2**; also see **Supplementary Figure 2** and **Supplementary Data**).

**Table 1:** Cumulative and age-stratified avertable COVID-19 deaths (ACD) in Iran (and 95% confidence interval) based on the per capita vaccine roll-out rates from model countries.

| **Model country** | **Fully vaccinated\*** | **ACD <50 age groups** | **ACD >50 age groups** | **Cumulative ACD** |
| --- | --- | --- | --- | --- |
| Argentina | 78.6% | 3 200 (2 300, 3 400) | 9 200 (8 100, 10 100) | 12 600 (10 400, 13 300) |
| Bahrain | 86.2% | 12 200 (8 900, 12 900) | 63 200 (47 700, 70 100) | 75 300 (56 000, 83 000) |
| Bangladesh | 70.5% | -20 000 (-80 700, -5 800) | -32 800 (-108 800, -11 700) | -52 800 (-189 500, -17 400) |
| Bolivia | 50.2% | -8 400 (-37 800, -1 800) | 1 800 (-21 600, 5 500) | -6 600 (-59 400, 3 600) |
| Montenegro | 46.9% | -9 100 (-42 500, -1 800) | 33 600 (32 800, 40 700) | 31 000 (-8 900, 31 600) |
| Nepal | 66.2% | -12 400 (-50 300, -3 600) | 7 200 (-10 300, 8 900) | -5 200 (-60 600, 5 300) |
| Sri Lanka | 70.0% | 2 300 (1 700, 2 500) | 21 800 (17 200, 21 900) | 24 200 (18 900, 24 400) |
| Turkey | 64.7% | 3 100 (2 100, 3 800) | 46 200 (35 100, 51 500) | 50 000 (38 100, 53 500) |

\*This is based on the counterfactual rescaled vaccination rate of the model countries for the Iranian population by the end of the study period (2022-04-20). The percentage of fully vaccinated individuals in Iran by the end of this period was 67.5%.

**Table 2:**
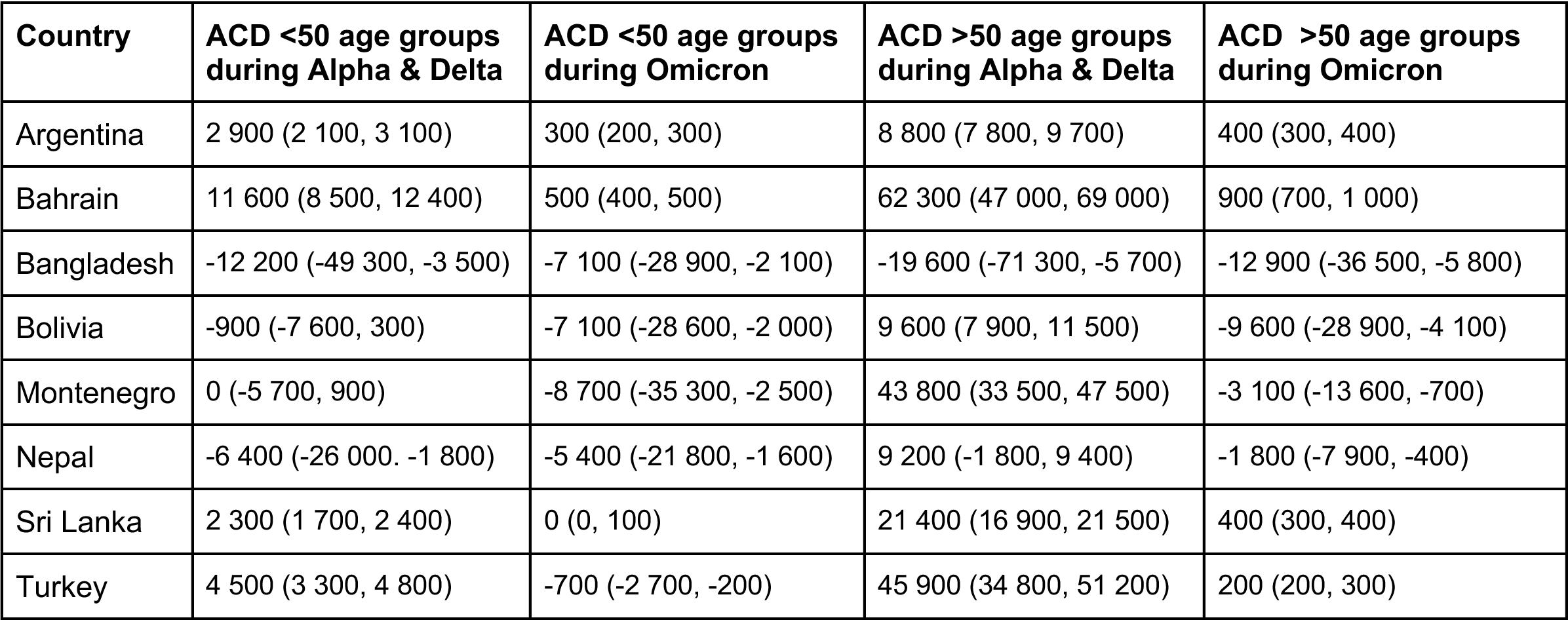
Age-stratified avertable COVID-19 deaths (ACD) in Iran based on the per capita vaccine roll-out rates from model countries at different stages of the epidemic in Iran. This includes ACDs during the Alpha and Delta waves (from 2021-02-19 to 2021-12-22) and the Omicron BA.1 wave (from 2022-01-05 to 2022-04-20).

We also found that following either Montenegro’s or Bolivia’s per capita roll-out rates, Iran could have averted many more deaths in the older age groups despite having a 17-20% lower percentage of fully vaccinated individuals (see **Table 1**). This is because faster roll-out rates in these countries would have enabled many more individuals to be protected in the >50 age groups during the Alpha and Delta waves. However, due to their lower overall percentage of fully vaccinated individuals, as many as 8,700 (95% CI: 2,000 - 35,300) more individuals in the <50 age groups could have lost their lives during the Omicron BA.1 wave (see **Table 2**). The reason for the wide uncertainty in the number of ACDs for these age groups is because if the vaccine effectiveness against death were very high (up to 90% protective against deaths), then having a lower coverage in younger age groups could have ended with many more deaths during the Omicron BA.1 wave (see **Methods** and **Table 2**).

To further investigate the impact of faster vaccine roll-out rates on ACDs, we also compared Bolivia’s and Nepal’s counterfactual vaccine roll-out rates for Iran. While both countries had similar overall outcomes in terms of cumulative ACDs by mid-April 2022 (see **Table 1**), following Nepal’s rates, Iran could have had as many as 6,400 (1,800 - 26,000) more deaths during Alpha and Delta waves in younger age groups. On the other hand, following Bolivia’s rates, Iran would have had 900 (300 - 7,600) more deaths in the same period and for the same age groups (see **Table 2**) despite having 16% lower overall coverage. This is also the same reason why Iran performed much worse than many other LMICs and UMICs with similar percentages of fully vaccinated individuals because during these two waves, particularly Delta, where the country had its highest per capita death rates (see **Supplementary Figure 2**), it could have averted many more deaths if it were to vaccinate the population earlier.

Finally, we also compared Iran’s vaccination roll-out against Bahrain, a country that had one of the fastest and most effective vaccination campaigns in the Eastern Mediterranean region [23], which can, therefore, provide a reasonable upper bound for the maximum number of ACDs in Iran. We found that following Bahrain’s model, a total of 75,300 (56,000 - 83,000) deaths could have been averted in Iran during the pandemic (**Table 1**). Majority of these ACDs would have happened in the >50 age groups during the Alpha and Delta waves from mid February to late December 2021 (**Table 2**; see also **Supplementary Data**).

## Discussion

In this study, we developed a quantitative framework to retrospectively estimate the number of COVID-19 vaccine-preventable deaths in a country by using per-capita vaccine roll-out rates from other countries as a basis for comparison. Iran was selected as a case study due to the delayed start of its national vaccination campaign which has been suggested to have contributed to a higher mortality and disease burden [18, 24, 25]. Our framework provides a simple and rapid assessment to gauge the impact of relative vaccination coverage, pace, and timing from different countries on avertable deaths. The method can also aid future decisions aimed at reducing the burden of infectious diseases under different vaccination strategies.

Our findings revealed that had Iran followed the per capita roll-out rates of certain model countries, it could have potentially averted a significantly higher number of deaths, particularly during the first nine months of 2021, as a result of faster COVID-19 vaccine roll-out rates. The impact of faster roll-outs would have been far greater in averting deaths in the older age groups due to their elevated age-stratified infection fatality rate [26, 27] while the impact of higher overall coverage would have been able to avert more deaths in the younger age groups. For instance, our results indicated that Iran could have saved nearly 31,000 more lives if it had followed the fast roll-out rates of Montenegro despite having a 20% lower overall coverage. This is because many more lives could have been saved in the older age groups during the Alpha and Delta waves despite having more deaths in the younger age groups during the Omicron BA.1 wave in Iran.

Comparisons with Bahrain, a country with a highly effective vaccination campaign, demonstrated that nearly 75,000 more deaths could have been averted in Iran by following a similar model. These findings showed the potential benefits of early vaccination start dates, but more importantly faster roll-out rates in reducing the disease burden. All of our estimates for ACDs should be treated as a lower bound for the true number of ACDs because we did not account for the indirect impact of vaccination on reducing deaths by lowering the transmission rates which would in turn reduce the total number of avertable deaths even further. Incorporating these effects into a full transmission dynamic model would likely depict a different trajectory for the epidemic curve in a population with higher vaccination rates. This is because early vaccination of more individuals would delay infections and potentially alter the dynamics of the epidemic by changing the patterns of immunity acquired through vaccination and infection [10]. However, accurate parameterisation of such a model would require taking into account other factors such as infection history, waning immunity, and the varied effectiveness of different vaccine types on reducing deaths for which the underlying data for Iran was lacking. Instead, we developed a fast and simple framework to estimate ACDs based on excess mortality data which is readily available for most countries around the world [28, 29].

Iran’s Ministry of Health pursued various strategies to increase its COVID-19 vaccine supply, including importations from Russia, India, and China, purchasement through the COVAX, and collaboration with Cuba for the development of the Soberana 02 and Soberana Plus vaccines, known as PastoCovac and PastoCovac Plus, respectively, in Iran. However, the prioritisation of domestic vaccine production, particularly the BIV1-CovIran vaccine from Barkat Pharmaceutical group which did not scale up in time, significantly delayed the vaccination roll-out in Iran. This was compounded by the concerns around lack of transparency in financial disclosures of researchers involved in the development of the Barekat vaccine [30, 31] and lack of data in support of its safety and efficacy in phase 3 clinical trials which as of yet has not been published despite receiving emergency use authorisation from the Iranian FDA almost two years ago [18].

In early-to mid-2021, Iran missed key opportunities to secure more vaccines. At the time, the policy of the Iranian Ministry of Health was not in favour of making prepayments for the purchase of vaccines from other countries but to instead focus on investing in local infrastructure to produce home-grown vaccines [32]. As a result, Iran missed an opportunity to secure high-enough vaccine doses through prepayments. Another missed opportunity was related to the decision by Iran’s Supreme Leader to ban vaccine imports from the US and UK. At the time of the announcement on 8 January 2021, Iran’ Red Crescent organisation, cancelled the importation of 150,000 doses of the donated Pfizer-BioNTech vaccine [18]. This alone could have protected up to 75,000 individuals (under a double dose vaccination schedule) and prevented thousands of deaths had they been administered to the elderly and most vulnerable individuals in time. Furthermore, given that mRNA vaccines elicit stronger humoral immunity relative to inactivated virus vaccines [33–35] which were widely used in Iran, it is plausible that they would have prevented more infections and deaths if they were more widely used or in combination with inactivated virus vaccines as heterologous vaccine regimens [36].

Another example of missed opportunities in Iran’s COVID-19 vaccination programme was the decision to not participate in phase 3 clinical trials of inactivated virus vaccines. Iran’s Ministry of Health had a policy to only participate in phase 3 trials of vaccines if the manufacturer makes a commitment to jointly collaborate with Iran in the technology transfer and production of vaccines. However, such an agreement was only reached with the Finley Institute of Cuba which ran its phase 3 trials in Iran [37, 38]. Nevertheless, this vaccine was not granted emergency use authorisation until much later and without any large importations from Cuba or local production in Iran, making up only 1.7% of all administered vaccine doses in Iran (**Supplementary Table 2**).

In contrast, some of the model countries included in this study such as Argentina, Bahrain, and Turkey all participated in trials for inactivated virus vaccines and were able to secure more doses at an earlier date compared to Iran. Bangladesh, on the other hand, did not participate in any phase 3 trials [39] and Nepal’s vaccination roll-out was in part affected by the halting of vaccine exports from India which the country relied on in 2021 [40]. By contrast, Montenegro did not participate in any clinical trials, but the rapid roll-out of vaccines in this country was facilitated by its relatively smaller size which made the deployment of vaccines easier [41].

## Conclusion

In conclusion, this study provided a quantitative framework to compare the performance of different countries based on their timing, pace, and coverage of vaccination on avertable COVID-19 deaths. It demonstrated the importance of faster roll-out rates on further reducing COVID-19 deaths and provided a means to find effective vaccination strategies in managing infectious disease epidemics based on country-level comparisons. Iran’s experience with COVID-19 vaccination also offers important lessons for public health policy decisions in future. The delayed start of their vaccination campaign as a result of the ban on vaccine imports from certain countries, lack of prioritisation on securing early vaccine doses, and over-reliance on the development and punctual delivery of home-grown vaccines in a period of public health emergency all had major public health implications that ultimately contributed to the lower number of vaccine-preventable deaths in Iran.

## Methods

We collected the number of fully vaccinated individuals from Our World in Data [23] and the WHO dashboard [42]. We obtained the economic status of each country from the World Bank income group in 2020 [43], a year prior to the start of vaccination campaigns in most countries. Our candidate model countries were selected based on having comparable income levels and predominant vaccine types to Iran, particularly those that predominantly use inactivated virus vaccines such as BBIBP-CorV vaccine of Sinopharm, as documented in the UNICEF COVID-19 Vaccine Market Dashboard [3, 44]. Since Iran was classed as UMIC before the commencement of vaccine distribution up to 2020 and LMIC from 2021 onwards, we included candidate model countries both from LMIC and UMIC with comparable income levels [43, 45, 46]. These countries include Argentina (UMIC), Bangladesh (LMIC), Bolivia (LMIC), Montenegro (UMIC), Nepal (LMIC), Sri Lanka (LMIC), and Turkey (UMIC). We also included Bahrain (HIC) as an upper bound benchmark for vaccine coverage relative to Iran.

We assumed that the vaccination roll-out in all countries would follow the same pattern as Iran’s where the older age groups and those at risk would be vaccinated first in a descending age order [24]. Since we only have information on the roll-out start dates for the first dose in each age group, we assumed that the time difference in receiving the full dose (two doses for most vaccines, one or three for a few other manufacturers) between each consecutive age group is the same as the time difference for the first dose (see **Supplementary Table 3**).

To calculate the net difference in the number of fully vaccinated individuals between Iran and modelled countries over time, we first took the per capita daily number of fully vaccinated individuals in model country *M*, *r_M_*(*t*) *= n_M_*(*t*)*/*P*_M_*, where *n*(*t*) is the daily number of fully vaccinated people and *P* is the total population size of the country. The net difference in the daily number of fully vaccinated individuals in age group *i*, Δ*n*^(*i*)^(*t*), in Iran relative to the vaccine roll-out rate of model country *M* then becomes:

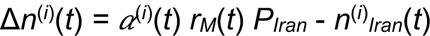

where *α*^(*i*)^(*t*) is the fraction of newly vaccinated individuals and *n*^(*i*)^*_Iran_* is the number of fully vaccinated individuals in age group *i* in Iran at time *t*.

The number of avertable deaths in age group *i* as a result of excess vaccine roll-out rate of model country *M* becomes:

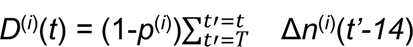

where *p*^(*i*)^ is the vaccine effectiveness against death in age group *i* and *T* is the date at which the first person in age group *i* is fully vaccinated. We assumed that fully vaccinated individuals become protected against deaths from COVID-19 two weeks after receiving the full dose and that the effectiveness of the vaccine against death is identical across all inactivated virus vaccines and remains the same over time such that *p*^(*i*)^*=* 0.923 (95% confidence interval: 0.672 - 0.982) for *i* > 60 years age groups and *p*^(*i*)^*=*0.801 (95% confidence interval: 0.611 - 0.898) for *i* ≤ 60 years age groups [47].

We used a previously published model for calculating the number of COVID-19 deaths in Iran using excess mortality data [24] across all age groups eligible to receive vaccination in Iran which includes 16 age groups *i ∈* {5-9,10-14, …,75-79, 80+} years old. Our analysis covers the period from the start of the pandemic up to 2022-04-20 as the association between excess mortality and reported COVID-19 deaths weakens after the end of the Omicron BA.1/2 wave (see **Supplementary Figure 2**).

## Data Availability

All data produced in this article are available online at https://ourworldindata.org/ and https://covid19.who.int/

## Acknowledgements

M.G. would like to thank Ariel Karlinsky and the Center for Interdisciplinary Data Science Research (CIDR) at the Hebrew University of Jerusalem for the analysis and sharing of excess mortality data.

## Conflicts on interest

None.

## Supplementary Tables

**Supplementary Table 1:** A timeline of vaccine imports to Iran from 02/02/2021 to 16/11/2021.

| Item No. | Vaccine type - originating country - cargo # | Date of entry | Quantity |
| --- | --- | --- | --- |
| 1 | Sputnik V - Russia - 1 | 03/02/2021 | 20,000 |
| 2 | Sputnik V - Russia - 2 | 13/02/2021 | 100,000 |
| 3 | Sinopharm - China - 1 | 28/02/2021 | 250,000 |
| 4 | Sputnik V - Russia - 2 | 04/03/2021 | 200,000 |
| 5 | Bharat - India - 1 | 10/03/2021 | 125,000 |
| 6 | Sputnik V - Russia - 4 | 25/03/2021 | 100,000 |
| 7 | Sputnik V - Russia - 5 | 01/04/2021 | 100,000 |
| 8 | AstraZeneca - South Korea - 1 | 05/04/2021 | 700,800 |
| 9 | Sinopharm - China - 2 | 15/04/2021 | 400,000 |
| 10 | Sputnik V - Russia - 6 | 22/04/2021 | 100,000 |
| 11 | Sputnik V - Russia - 7 | 29/04/2021 | 100,000 |
| 12 | Sinopharm - China - 3 | 02/05/2021 | 1,000,000 |
| 13 | Sinopharm - China - 4 | 16/05/2021 | 1,000,000 |
| 14 | AstraZeneca - Italy - 1 | 16/05/2021 | 1,452,000 |
| 15 | Sputnik V - Russia - 8 | 03/06/2021 | 100,000 |
| 16 | Sputnik V - Russia - 9 | 10/06/2021 | 100,000 |
| 17 | Sinopharm - China - 5 | 17/06/2021 | 1,000,000 |
| 18 | Sinopharm - China - 6 | 24/06/2021 | 1,000,000 |
| 19 | Sinopharm - China - 7 | 08/07/2021 | 1,056,000 |
| 20 | Sinopharm - China - 8 | 10/07/2021 | 500,000 |
| 21 | Sinopharm - China - 9 | 11/07/2021 | 444,000 |
| 22 | Sinopharm - China - 10 | 15/07/2021 | 1,142,800 |
| 23 | Sinopharm - China - 11 | 18/07/2021 | 1,130,600 |
| 24 | Sinopharm - China - 12 | 22/07/2021 | 1,130,400 |
| 25 | AstraZeneca - Japan - 1 | 23/07/2021 | 1,087,570 |
| 26 | Sinopharm - China - 13 | 25/07/2021 | 1,095,200 |
| 27 | AstraZeneca - Russia (private sector) - 1 | 28/07/2021 | 315,000 |
| 28 | Sputnik V - Russia - 10 | 29/07/2021 | 100,000 |
| 29 | AstraZeneca - Japan - 2 | 30/07/2021 | 1,098,300 |
| 30 | AstraZeneca - Japan - 3 | 30/07/2021 | 725,940 |
| 31 | Sinopharm - China - 14 | 01/08/2021 | 1,120,000 |
| 32 | Sinopharm - China - 15 | 05/08/2021 | 1,110,000 |
| 33 | Sinopharm - China - 16 | 08/08/2021 | 1,110,000 |
| 34 | Sinopharm - China - 17 | 12/08/2021 | 1,110,000 |
| 35 | Sinopharm - China - 18 | 12/08/2021 | 500,000 |
| 36 | Sinopharm - China - 19 | 15/08/2021 | 1,110,000 |
| 37 | Sinopharm - China - 20 | 17/08/2021 | 700,000 |
| 38 | Sinopharm - China - 21 | 19/08/2021 | 1,110,000 |
| 39 | Sputnik V - Serbia - 11 | 20/08/2021 | 25,000 |
| 40 | Sinopharm - China - 22 | 22/08/2021 | 1,110,000 |
| 41 | Sinopharm - China - 23 | 26/08/2021 | 2,220,000 |
| 42 | AstraZeneca - China - 1 | 02/09/2021 | 1,449,600 |
| 43 | Sinopharm - China - 24 | 02/09/2021 | 179,596 |
| 44 | Sinopharm - China - 25 | 03/09/2021 | 4,115,088 |
| 45 | AstraZeneca - Russia (private sector) - 2 | 04/09/2021 | 648,000 |
| 46 | Sinopharm - China - 26 | 05/09/2021 | 5,000,000 |
| 47 | Sinopharm - China - 27 | 07/09/2021 | 897,014 |
| 48 | AstraZeneca - Austria - 1 | 08/09/2021 | 500,000 |
| 49 | Sinopharm - China - 28 | 09/09/2021 | 5,000,000 |
| 50 | Sinopharm - China - 29 | 11/09/2021 | 3,196,800 |
| 51 | Sinopharm - China - 30 | 16/09/2021 | 5,000,000 |
| 52 | Sputnik V - Russia - 12 | 16/09/2025 | 300,000 |
| 53 | AstraZeneca - Austria - 2 | 16/09/2021 | 150,000 |
| 54 | Sinopharm - China - 31 | 17/09/2021 | 3,000,000 |
| 55 | Sinopharm - China - 32 | 21/09/2021 | 2,000,000 |
| 56 | Sinopharm - China - 33 | 23/09/2021 | 5,000,000 |
| 57 | Sinopharm - China - 34 | 26/09/2021 | 6,000,000 |
| 58 | Sputnik V - Serbia - 13 | 27/09/2021 | 25,000 |
| 59 | Sinopharm - China - 35 | 28/09/2021 | 2,000,000 |
| 60 | Sinopharm - China - 36 | 30/09/2021 | 6,000,000 |
| 61 | AstraZeneca - Greece - 1 | 01/10/2021 | 150,000 |
| 62 | AstraZeneca - Austria - 3 | 03/10/2021 | 350,000 |
| 63 | Bharat - India - 2 | 09/10/2021 | 1,000,000 |
| 64 | Sinopharm - China - 37 | 10/10/2021 | 6,000,000 |
| 65 | Sinopharm - China - 38 | 12/10/2021 | 6,000,000 |
| 66 | Sinopharm - China - 39 | 14/10/2021 | 6,000,000 |
| 67 | Sputnik V - Serbia - 14 | 14/10/2021 | 400,000 |
| 68 | AstraZeneca - Poland -1 | 16/10/2021 | 1,000,000 |
| 69 | Sinopharm - China - 40 | 17/10/2021 | 6,000,000 |
| 70 | AstraZeneca - Netherlands (gift from Italy) - 1 | 18/10/2021 | 1,442,000 |
| 71 | Sinopharm - China - 41 | 19/10/2021 | 6,000,000 |
| 72 | Sinopharm - China - 42 | 21/10/2021 | 6,000,000 |
| 73 | Sputnik V - Russia - 15 | 21/10/2021 | 777,600 |
| 74 | Sinopharm - China - 43 | 24/10/2021 | 6,000,000 |
| 75 | Sinopharm - China - 44 | 26/10/2021 | 6,000,000 |
| 76 | AstraZeneca - South Korea - 2 | 27/10/2021 | 1,000,000 |
| 77 | Sinopharm - China - 45 | 28/10/2021 | 6,000,000 |
| 78 | Sputnik V - Russia - 16 | 29/10/2021 | 766,000 |
| 79 | Sinopharm - China - 46 | 31/10/2021 | 6,000,000 |
| 80 | Sinopharm - China - 47 | 02/11/2021 | 148,800 |
| 81 | Sputnik V - Russia - 17 | 04/11/2021 | 777,600 |
| 82 | Sinopharm - China - 48 | 15/11/2021 | 610,422 |
| 83 | Sinopharm - China - 49 | 16/11/2021 | 407,490 |
| 84 | Sinopharm - China - 50 | 16/11/2021 | 593,688 |
| 85 | AstraZeneca - Germany - 1 | 09/12/2021 | 131,000 |
| 86 | AstraZeneca - Netherlands - 2 | 09/12/2021 | 1,087,400 |
| 87 | Sinopharm - China - 51 | 16/11/2021 | 105,600 |
| Cargoes | 87 | Grand Total | 150,108,308 |

**Supplementary Table 2:** Total number of administered doses of COVID-19 vaccines in Iran per vaccine name by 29 May 2022 [48].

| **Vaccine Name** | **Acquisition type** | **Total administered doses** |
| --- | --- | --- |
| **Sputnik V** | Bilateral | 1,380,958 |
| **AstraZeneca** | COVAX + Bilateral | 14,044,541 |
| **Covaxine** | Bilateral | 254,520 |
| **Sinopharm (BBIBP-CorV)** | COVAX + Bilateral + Donation | 120,878,057 |
| **COVIran Barekat** | Local | 9,037,924 |
| **PastoCoVac** | Local | 2,485,211 |
| **SpikoGene** | Local | 1,091,761 |
| **Fakhravac** | Local | 65,200 |
| **Razi CovPars** | Local | 119,676 |
| **Noora** | Local | 10,405 |

**Supplementary Table 3:** Iran’s vaccine roll-out dates per age-group [24].

| **Age group** | **Roll-out date** | **Days since roll-out of first age group** |
| --- | --- | --- |
| +80 | 2021-05-08 | 0 |
| +75 | 2021-05-18 | 10 |
| +70 | 2021-05-22 | 14 |
| +65 | 2021-07-11 | 64 |
| +60 | 2021-07-14 | 67 |
| +55 | 2021-07-24 | 77 |
| +50 | 2021-07-30 | 83 |
| <50 | 2021-08-03 | 87 |

## Supplementary Figures

**Supplementary Figure 1:**
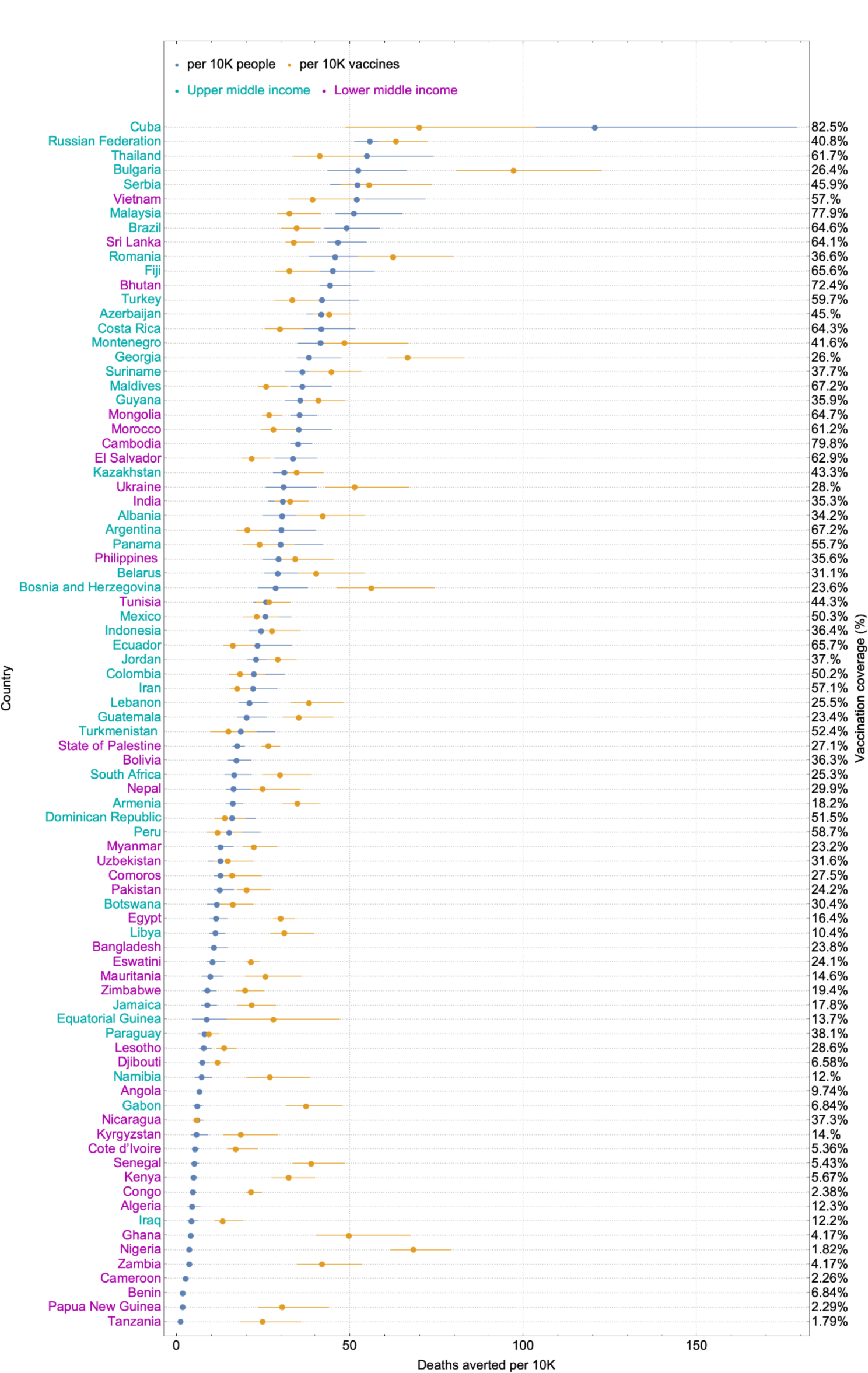
Deaths averted by vaccination for upper and lower middle income countries, including Iran, up to 2021-12-08. Data on deaths averted per person and per vaccine are downloaded from https://github.com/mrc-ide/covid-vaccine-impact-orderly/releases/download/v1.0.1/excess_mortality_summary_table.csv [3]. The economic status of each country is based on the World Bank income groups in 2020 [43].

**Supplementary Figure 2:**
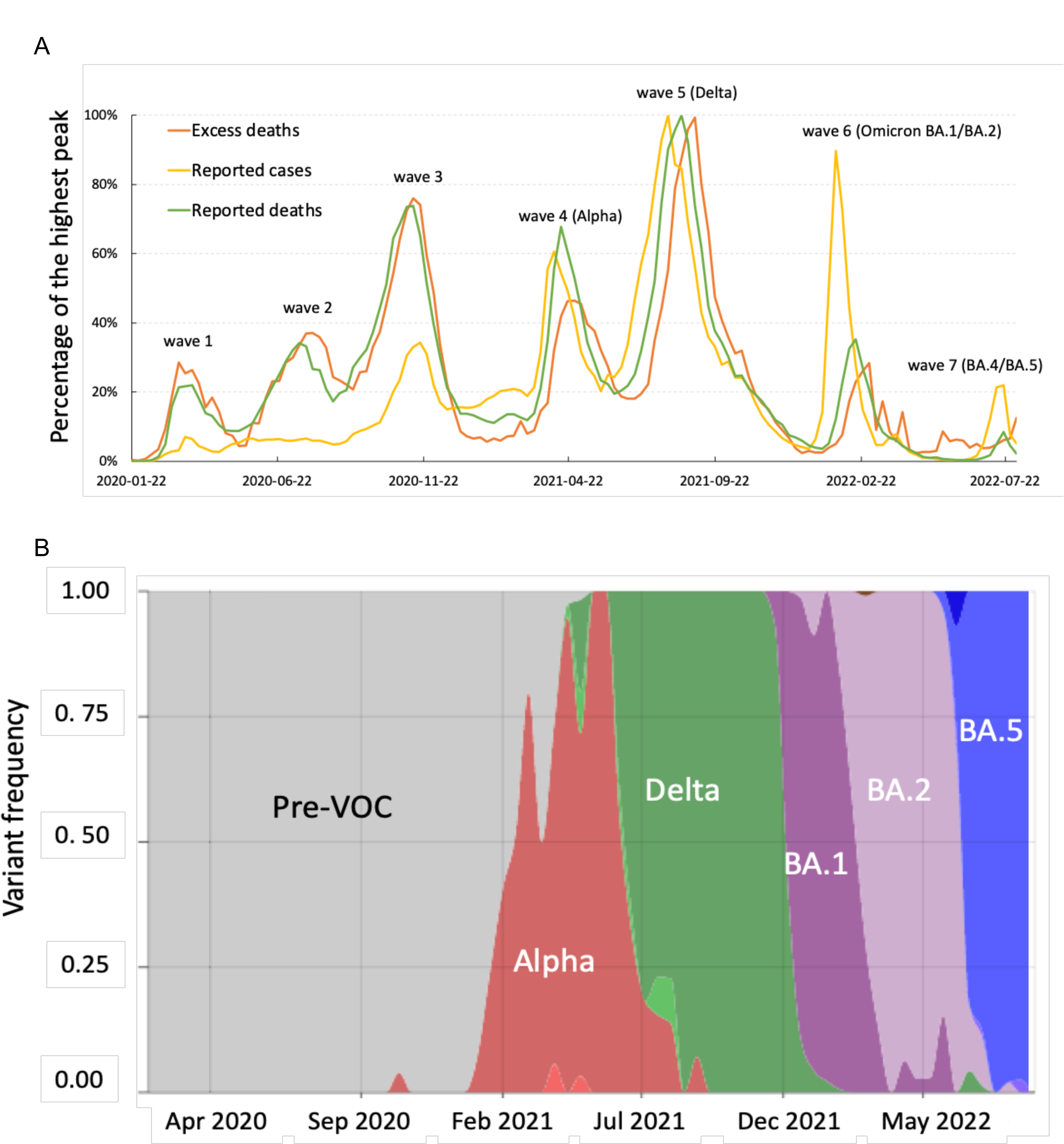
Association between reported cases, deaths, and excess deaths over time and with respect to variants of SARS-CoV-2. (A) Reported cases, deaths, and excess deaths (as quantified by the sum of excess deaths per age group) as percentage of highest peak (Delta) in Iran over time. (B) Proportion of the total number of SARS-CoV-2 variants over time in Iran. Data obtained from [49].

## Supplementary Data

**Avertable COVID-19 deaths in Iran over time based on the per capita vaccine roll-out rates from model countries**. Top panel shows Iran’s weekly excess deaths (black) and counterfactual excess deaths (magenta) had it followed the vaccination rate for each model country. Shaded areas show the 95% confidence interval for the counterfactual excess deaths based on varying degree of vaccine effectiveness against deaths. Central panel shows the avertable deaths based on vaccination rates from a given model country. It shows the difference between weekly excess deaths and counterfactual excess deaths in the top panel. Bottom panel shows the percentage of excess vaccination for a given model country relative to Iran’s vaccination rates per age group. Shaded areas in green (red) show periods where there would have been more (less) vaccination had Iran followed the vaccination rates as the model country.

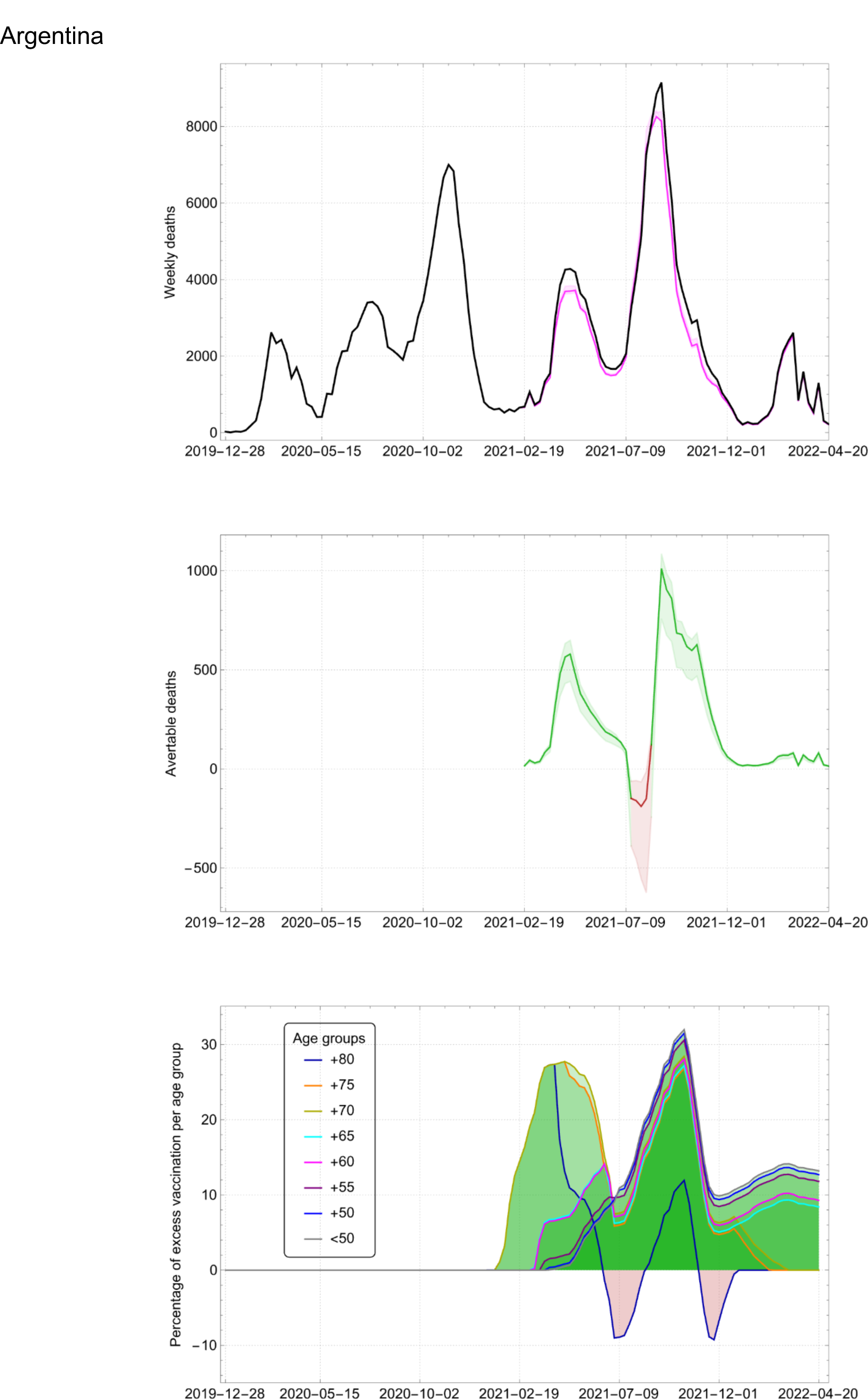

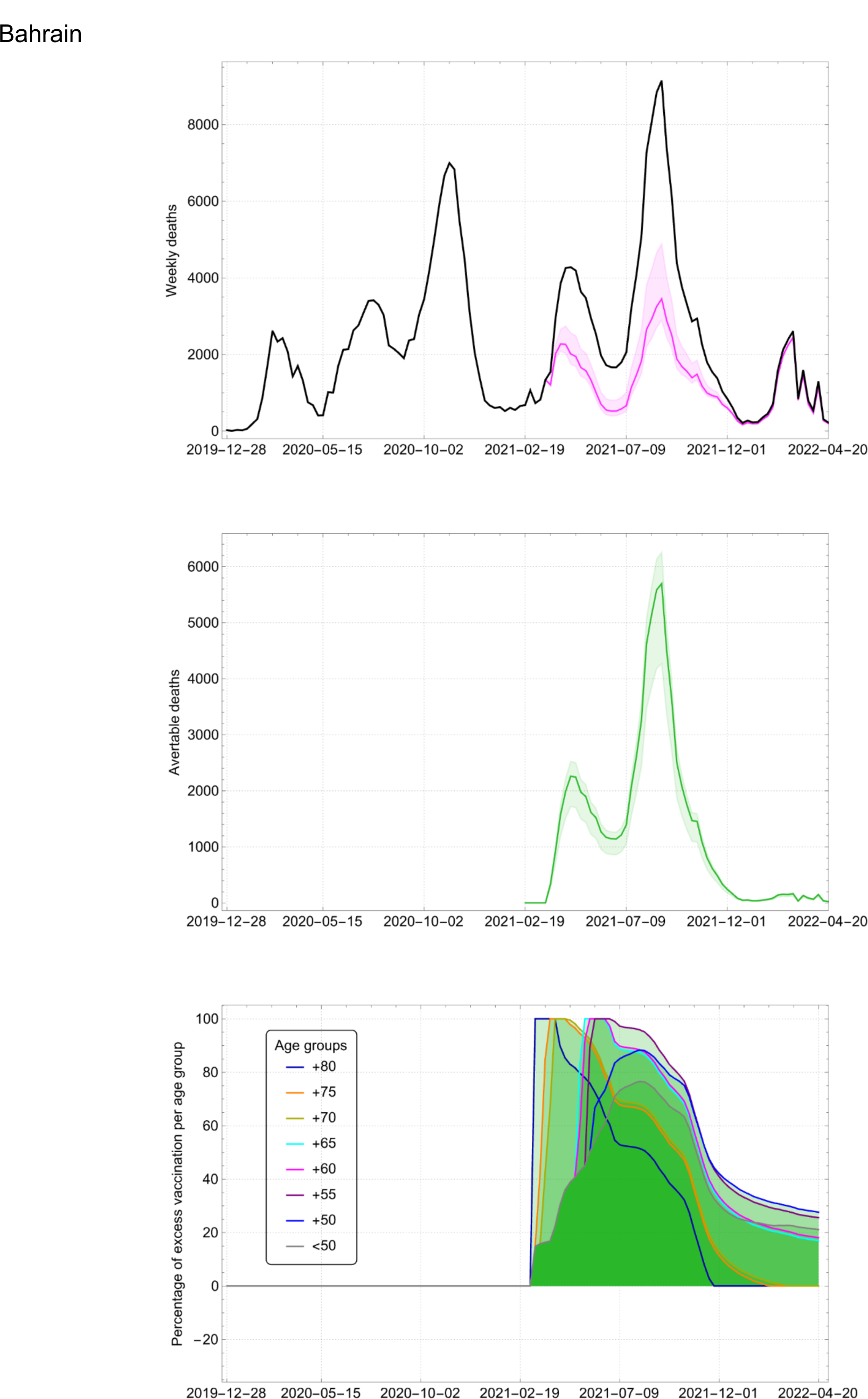

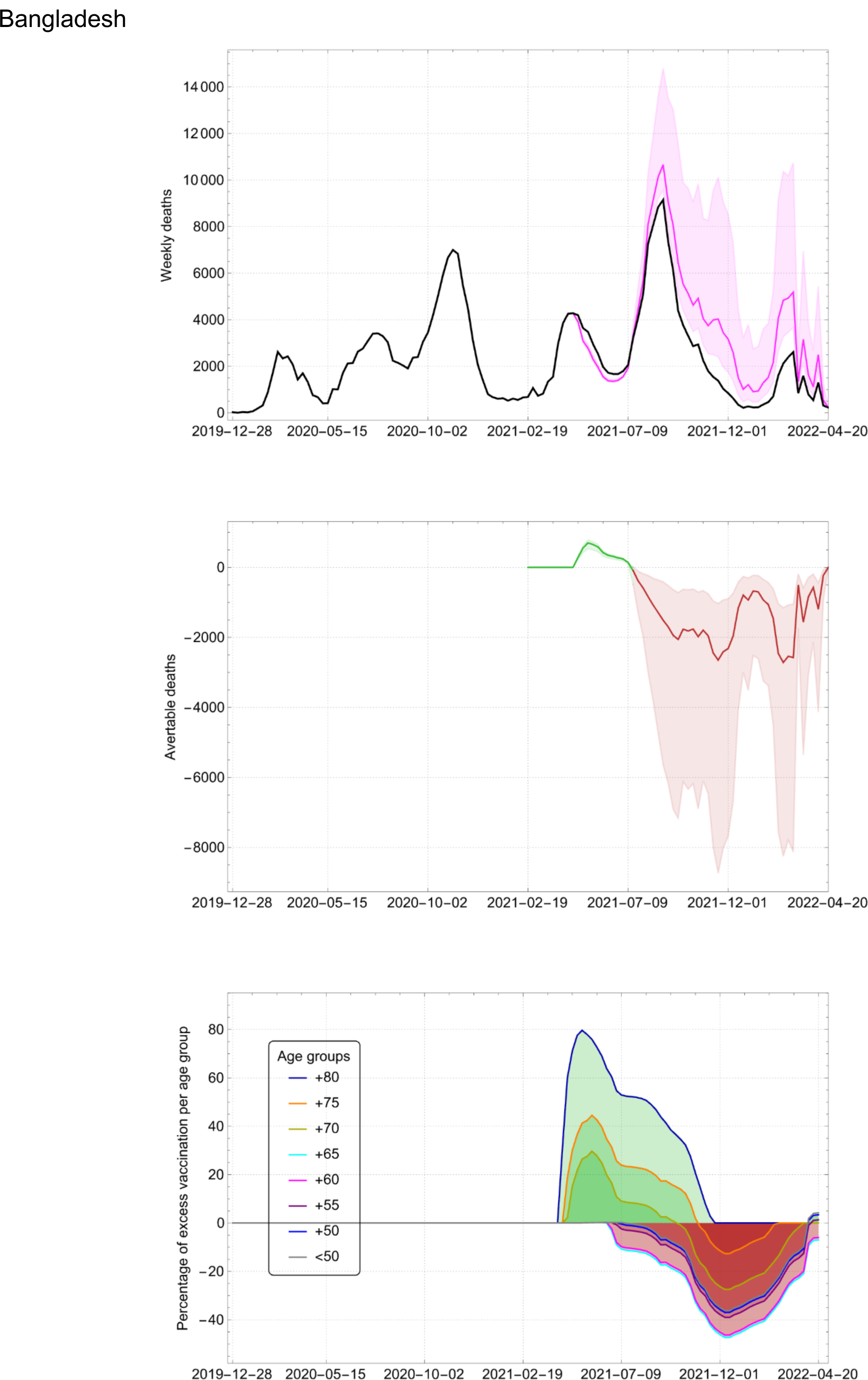

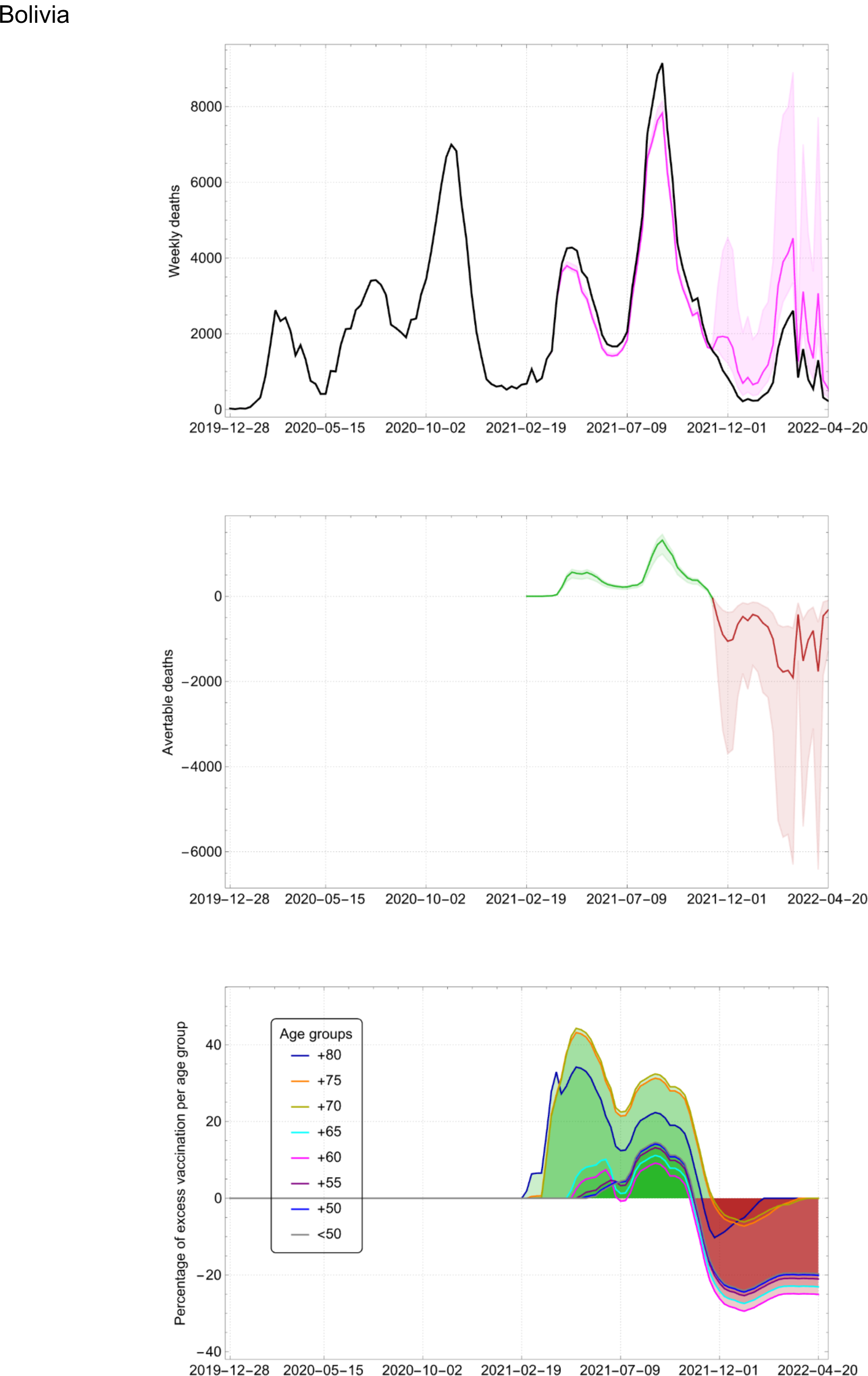

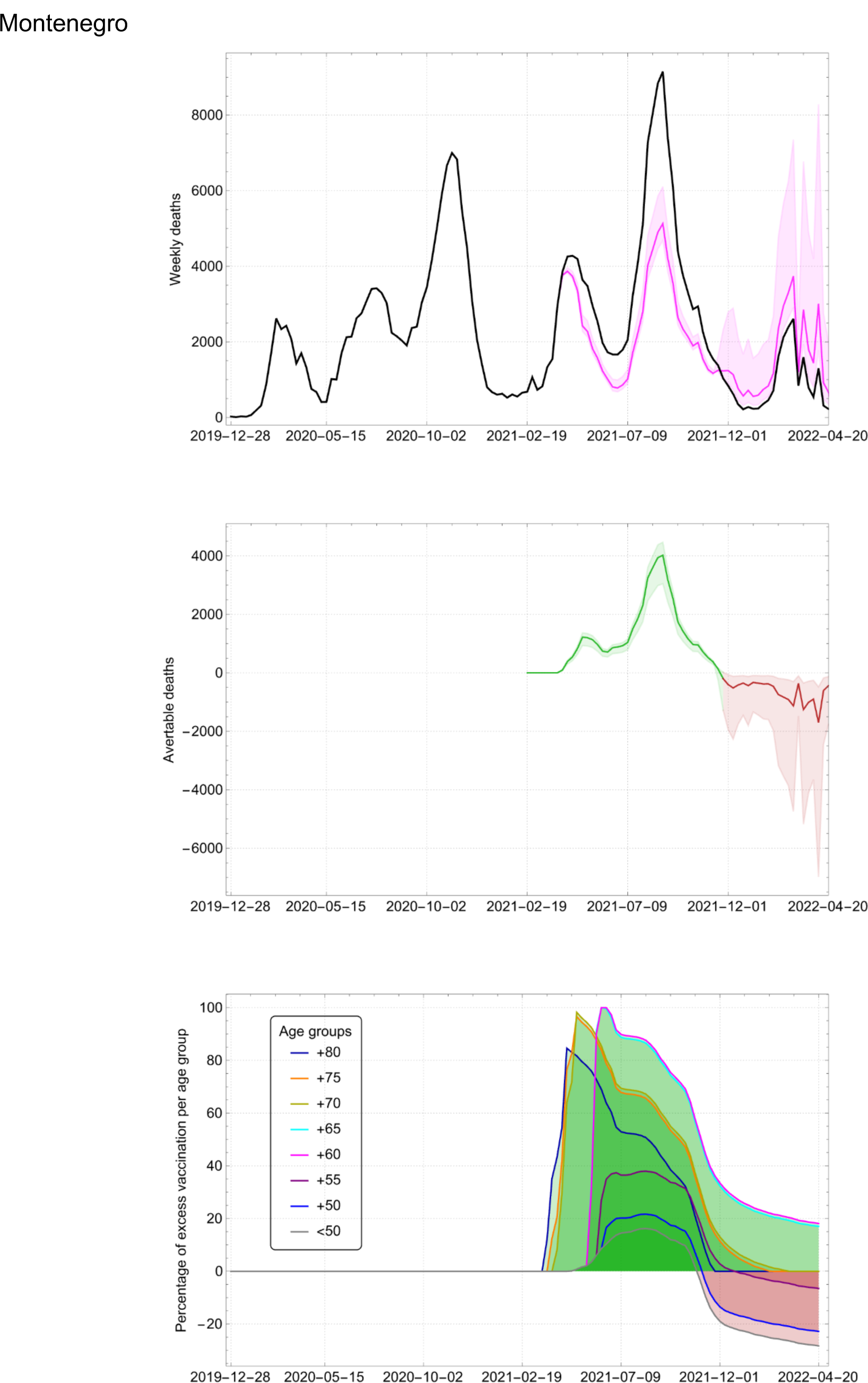

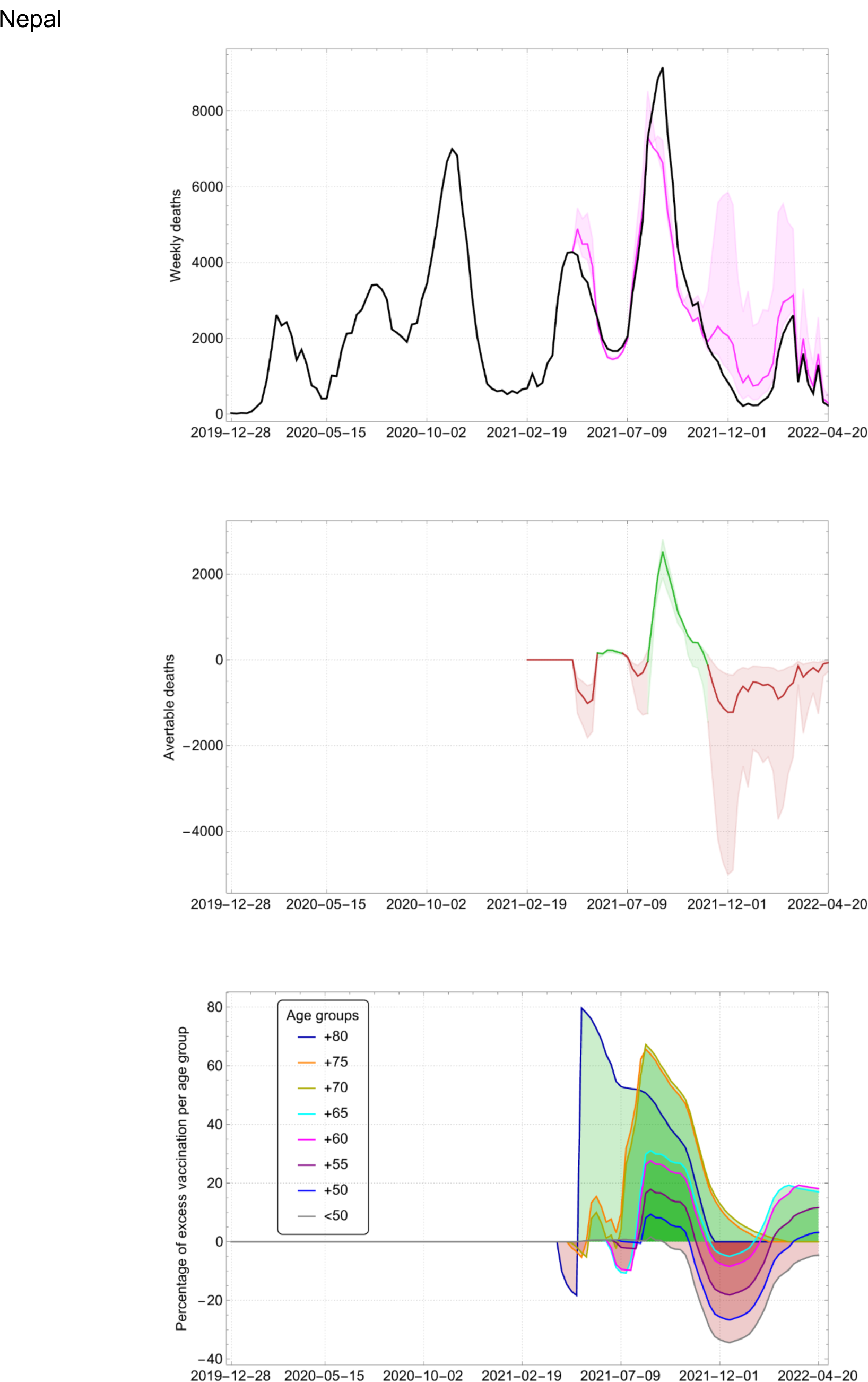

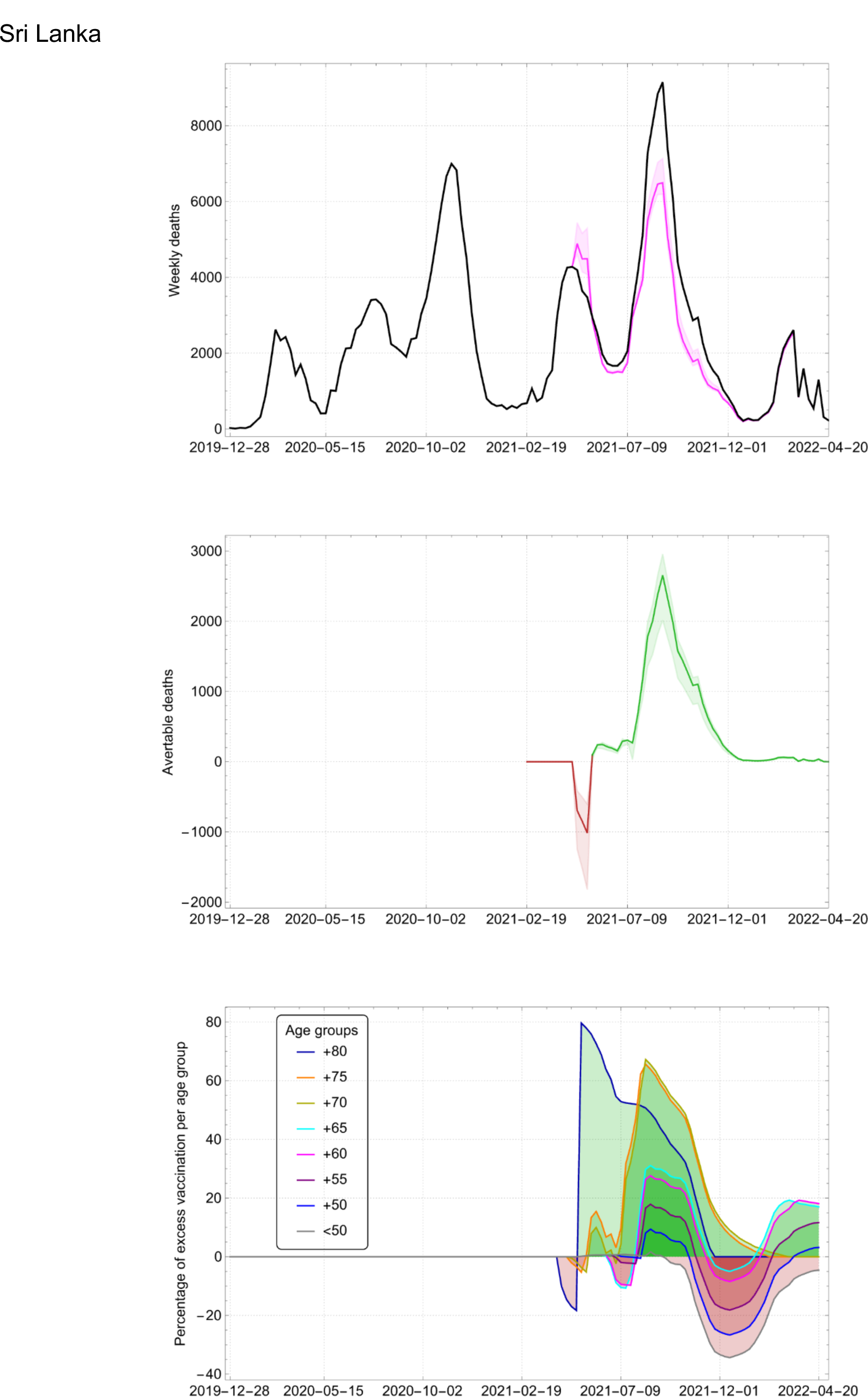

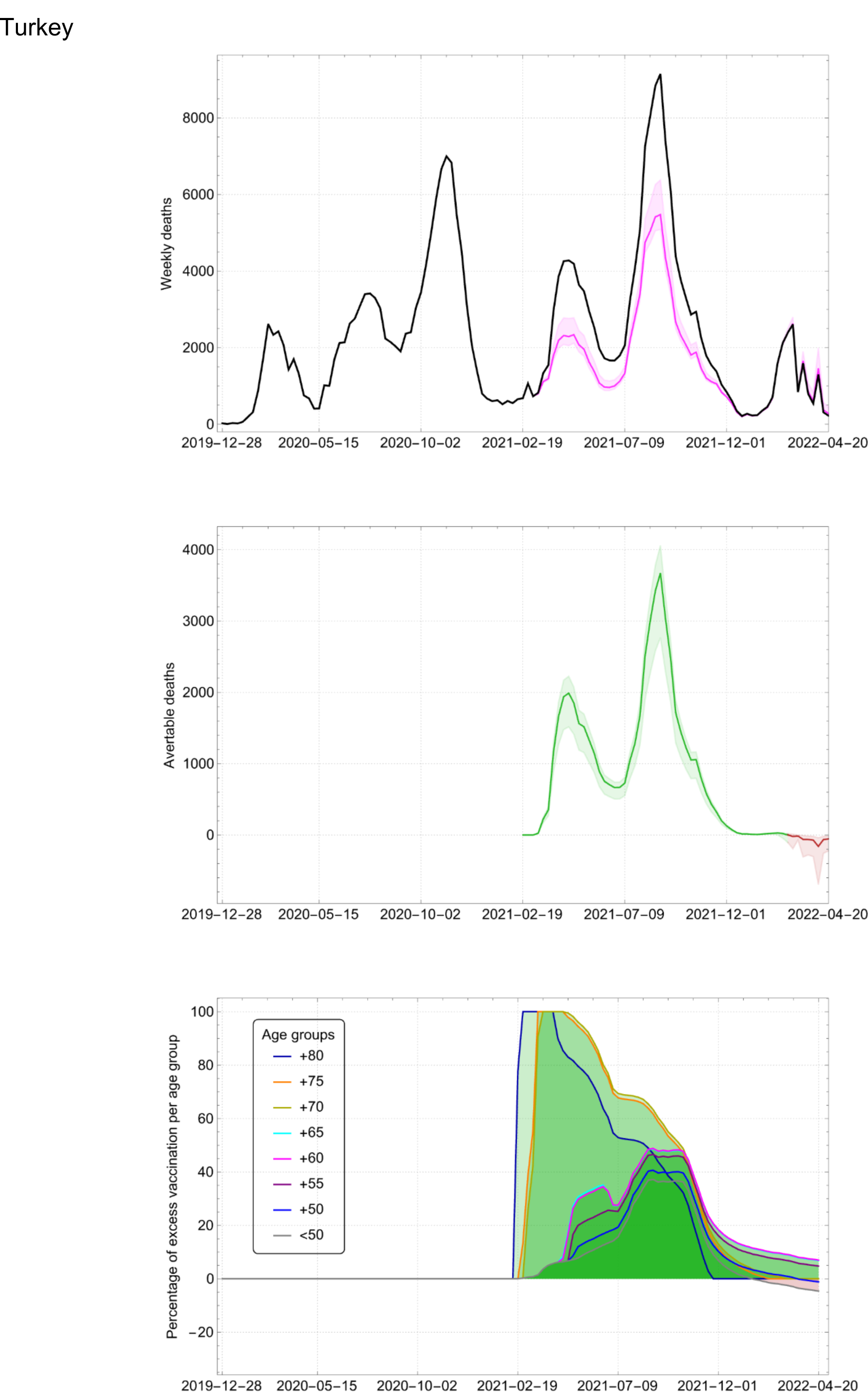

## Notes

### Competing Interest Statement

The authors have declared no competing interest.

### Funding Statement

This study did not receive any funding.

